# CovidArray: a microarray-based assay with high sensitivity for the detection of SARS-CoV-2 in nasopharyngeal swabs

**DOI:** 10.1101/2021.01.21.21250281

**Authors:** Francesco Damin, Silvia Galbiati, Stella Gagliardi, Cristina Cereda, Francesca Dragoni, Claudio Fenizia, Valeria Savasi, Laura Sola, Marcella Chiari

**Affiliations:** Istituto di Scienze e Tecnologie Chimiche “Giulio Natta” SCITEC CNR, Milan, Italy; Complications of Diabetes Units, Diabetes Research Institute, IRCCS San Raffaele Scientific Institute, Milan, Italy; Genomic and Post Genomic Unit, IRCCS Mondino Foundation, Pavia, Italy; Department of Biology and Biotechnology “L. Spallanzani”, University of Pavia, Pavia, Italy; Department of Pathophysiology and Transplantation, University of Milan, Milan, Italy; Unit of Obstetrics and Gynecology, L. Sacco Hospital ASST Fatebenefratelli Sacco; Department of Biomedical and Clinical Sciences, University of Milan, Milan, Italy

**Author notes:** **Corresponding Author** Francesco Damin. Istituto di Scienze e Tecnologie Chimiche “Giulio Natta” SCITEC CNR, Milan, Italy, Via Mario Bianco 9, 20131 Milano, Italy. F. Damin and S. Galbiati contributed equally to this work.

**Keywords:** SARS-CoV-2, microarray, RT-qPCR, microarray-based assay, Covid-19, molecular diagnostics

## Abstract

**Background:** A new coronavirus (SARS-CoV-2) caused the current Covid-19 epidemic. Reverse transcription-quantitative polymerase chain reaction (RT-qPCR) is used as the gold standard for clinical detection of SARS-CoV-2. Under ideal conditions RT-qPCR Covid-19 assays have analytical sensitivity and specificity greater than 95%. However, when the sample panel is enlarged including asymptomatic individuals, the sensitivity decreases and false-negative are reported. Moreover, RT-qPCR requires up to 3-6 hours with most of the time involved in RNA extraction from swab samples.

**Methods:** We introduce CovidArray, a microarray-based assay, to detect SARS-CoV-2 markers N1 and N2 in the nasopharyngeal swabs. The method is based on solid phase hybridization of fluorescently labelled amplicons upon RNA extraction and reverse transcription. This approach combines the physical-optical properties of the silicon substrate with the surface chemistry used to coat the substrate to obtain a diagnostic tool of great sensitivity. Furthermore, we used an innovative approach, RNAGEM, to extract and purify viral RNA in less than 15 minutes. To validate the CovidArray results, we exploited the high sensitivity of the droplet digital PCR (ddPCR) technique.

**Results:** We correctly assigned 12 nasopharyngeal swabs, previously analyzed by RT-qPCR. Thanks to the CovidArray sensitivity that matches that of the ddPCR, we were able to identify a false-negative sample.

**Conclusions:** CovidArray is the first DNA microarray-based assay to detect viral genes in the swabs. Its high sensitivity and the innovative viral RNA extraction by RNAGEM allows to reduce both the amount of false negative results and the total analysis time to about 2 hours.

## Introduction

In December 2019, an unexplained acute respiratory disease, Covid-19, emerged in Wuhan, China (1). It was immediately determined that the disease’s cause was a novel coronavirus named SARS-CoV-2 (2). Since virus identification and sequencing in early January 2020 (3), the primary approach for detecting viral RNA in respiratory specimens was the reverse transcription-quantitative polymerase chain reaction (RT-qPCR) (4,5). The RT-qPCR, initially used to confirm symptomatic patients’ diagnosis, was increasingly used to screen asymptomatic contacts and subjects at risk. Several RT-qPCR assays have been developed and recommended by the World Health Organization, the United States Centers for Disease Control and Prevention, the Chinese Center for Disease Control and Prevention, as well as by private companies (6,7). In the absence of specific therapeutic drugs for Covid-19, it is essential to detect the disease at early stage and immediately isolate the infected person from a healthy population. RT-qPCR Covid-19 assays have analytical sensitivity and specificity greater than 95% (8). However, this number refers to tests validated under ideal conditions with hospital samples containing viral loads higher than those from asymptomatic individuals. When the sample panel is enlarged to include asymptomatic individuals, the sensitivity decreases and false-negative rates between 2% and 33% (8) are reported. In the current emergency, individuals with Covid-19 that are not identified and quarantined represent a transmission vector for a larger amount of the population given the highly contagious nature of the virus. Failures in SARS-CoV-2 detection may be related to multiple preanalytical and analytical factors, such as lack of standardization for specimen collection, poor storage conditions before arrival in the laboratory and lack of specialized personnel for collection and analysis of the samples. Furthermore, poorly validated assays, contamination during the procedure, insufficient viral specimens or viral load contribute to results’ uncertainty. The variable viral load along the disease’ incubation period, or the presence of mutations and PCR inhibitors (9,10) also impact the diagnosis.

In addition, shortage of reagents is also greatly contributing to spread the disease by delaying the Covid-19 diagnostics and reducing the number of tests available. Many regions around the world have experienced a shortage of laboratory-based molecular-assay tests. Executing a test requires about 20 different reagents, consumables, and other pieces of equipment. Of those materials, major shortages have been reported in RNA-extraction kits (11).

We propose an approach to detect SARS-CoV-2 endowed with high sensitivity, accuracy, and multiplexing capability based on microarray technology, named CovidArray. The method is based on solid phase hybridization of fluorescently labelled amplicons upon RNA extraction, and reverse transcription. Essential features of this system, already successfully applied in liquid biopsy and prenatal diagnosis (12-14), are: i) the use of crystalline silicon chips coated by a 100 nm thermally grown silicon dioxide (SiO_2_) layer to enhance fluorescence signals (15), and ii) the efficient surface chemistry used to bind to the substrate the oligonucleotide capture probes specific to the virus genes (16). A three-dimensional coating made of copoly(DMA-NAS-MAPS), a copolymer known for its high binding capacity and low non-specific adsorption, was used (17). To validate the microarray-based assay results, we exploited the high sensitivity of the droplet digital PCR (ddPCR) technique. ddPCR, whose limit of detection (LOD) has been shown by several works to be significantly lower than that of qPCR (18,19), is based on the principles of limited dilution, end-point PCR, and Poisson statistics, with absolute quantification as its heart (20).

In this work, we followed the guidelines of the US Centers for Disease Control and Prevention (CDC) and the CDC 2019-Novel Coronavirus (2019-nCoV) RT-qPCR Diagnostic Panel (21) for the qualitative detection of RNA from SARS-CoV-2 in upper and lower respiratory specimens (such as nasopharyngeal or oropharyngeal swabs). The oligonucleotide primers and probes were selected from two virus nucleocapsid (N1 and N2) gene regions. An additional set of primers/probe to detect the human Ribonuclease P gene (*RPP30*) as a control of nucleic acid extraction in clinical specimens was also included in the panel. Many available commercial RT-qPCR kits employ multiplex system capable of detecting 2 or 3 different SARS-CoV-2 targets as well as an internal control. Similarly, the CovidArray assay is able to perform a multiplex detection of the markers N1, N2 and *RPP30*, but, compared to the commercial RT-qPCR kits, our system has the additional potential to differentiate SARS-CoV-2 from other viral and bacterial respiratory tract infections simply by adding new primers and capture probes to the same array.

Another key improvement introduced by CovidArray is the faster analysis time. An important feature of a diagnostic assay during an outbreak is the overall execution time since a fast method would allow expanding the analytical throughput. The standard methodology for SARS-CoV-2 detection requires from 3 to 6 hours to run a test (22) with most of the time involved in RNA extraction from swab samples, with CovidArray the time required is reduced to about 2 hours. Many commercial RNA extraction kits such as, for example, the Roche MagNA Pure 96 or the QIAGEN QIAcube kits have been validated for viral RNA extraction purpose (21). Although the RNA isolation kits are easy to use in automated instruments, it might be necessary to use alternative approaches to extraction to expand the analytical capability in the case of an epidemic. In this work, we used an innovative single-tube approach to extract the viral RNA, by employing RNAGEM, a straightforward temperature-driven enzymatic method to extract RNA, commercially available by MicroGEM (MicroGEM UK Ltd, Southampton). The main advantages of using this extraction methodology are: i) minimal pipetting steps (manual or automated) leading to less contamination, virtually no loss of RNA and reduced amount of plastic consumables (pipette tips, tubes, etc; also in shortage during this pandemic), ii) no need of using a harsh chemical which eliminates the washing steps, iii) no need for further purification of the RNA for accurate RT-qPCR and qPCR analysis and iv) single-tube workflow that provides purified RNA in 10 minutes, v) extraction is conducted using a common laboratory thermocycler allowing to extract up to 96 samples simultaneously, vi) the manual steps can be automated easily by using any liquid handler.

Moreover, here we demonstrated that combination of single-tube extraction by RNAGEM with highly sensitive multiplex microarray substrate with optimal properties allow to reduce the number of PCR cycles from 40 to 25 and to lead to an overall increase in accuracy and a reduction in analysis time.

## Materials and methods

### Materials and reagents

Copoly(DMA-NAS-MAPS) (MCP-4) was obtained by Lucidant Polymers Inc., Sunnyvale CA, USA. Ammonium sulphate ((NH_4_)_2_SO_4_), ethanolamine and 20X standard saline sodium citrate (SSC) solution (3 M sodium chloride, 0.3 M sodium citrate, pH 7.0), sodium dodecyl sulfate (SDS), were purchased from Sigma Aldrich (St. Louis, MO, USA). All the oligonucleotides were synthesized by Metabion International AG (Steinkirchen, Germany). Their sequences are reported in Supplemental Table1. Untreated silicon/silicon oxide chips with 100 nm thermal grown oxide (15 x 15 mm) were supplied by SVM, Silicon Valley Microelectronics Inc. (Santa Clara, CA, USA). Chips were pretreated using a HARRICK Plasma Cleaner, PDC-002 (Ithaca, NY, USA) connected to an oxygen line.

Spotting is perfomed using a SciFLEXARRAYER S12 (Scienion, Berlin, Germany). InnoScan 710 (Innopsys, Carbonne, France) was used to scan the hybridized chips. Data intensities were extracted with the Mapix software and the data analysis was performed for each experiment.

A detailed description of the samples collection, RT-qPCR, Reverse Transcription and ddPCR and 2019-CoV Plasmid Controls is provided in the Online Supplemental Data.

### RNA extraction for CovidArray analysis

The viral RNA was extracted by mixing 89.5 µL of the inactivated UTM (70°C for 1 h) containing a nasopharyngeal swab with 0.5 µL of RNAGEM enzyme (MicroGEM UK Ltd, Southampton) and 10 µL of 10X Blue Buffer. Subsequently, the extraction was conducted by incubating the reaction mix in a thermocycler at 75°C for 5 min and 95°C for 5 min.

### PCR conditions for microarray

The N1, N2 and *RPP30* sequences were amplified using the 5’-biotin forward and 5’-Cy3 labelled reverse primers reported in Supplemental Table 1.

The PCRs were performed in 20 µL of reactions containing 9 µL of cDNA previously diluted 1:20, 200 µM deoxynucleotide triphosphates, 10 mM Tris–HCl (pH 8.3), 50 mM KCl, 1.5 mM MgCl_2_, 1 U of DNA polymerase (FastStart Taq, Roche) and 10 pmoles of each primer.

Cycling conditions were as follows: 95 °C for 5 min; 25 cycles at 95°C for 30 s, 60°C for 30 s, 72°C for 30 s and finally 72°C for 10 min.

In addition to the single amplification, we optimized a triplex PCR amplification in which the primers used for the N1, N2 and *RPP30* amplification were mixed in the same PCR mixture.

The triplex PCR was performed in 20 µL of reactions containing 9 µL of diluted cDNA, 10 pmoles of each primer and 4 µL of 5X HOT FIREPol Blend Master Mix Ready to Load (Solis BioDyne).

Cycling conditions were as follows: 95 °C for 12 min; 25 cycles at 95°C for 20 s, 60°C for 30 s, 72°C for 30 s and finally 72°C for 7 min.

### Microarray preparation

We selected three oligonucleotide sequences from the US CDC 2019-Novel Coronavirus (2019-nCoV) Real-Time RT-qPCR Diagnostic Panel corresponding to the two specific probes for the regions N1 and N2 of the virus nucleocapsid gene and to the human *RPP30* gene. In addition, an oligonucleotide probe, not correlated to any viral or human sequences, was selected as negative hybridization control. Capture and control probes (reported in Supplemental Table 1), amino modified at the 5’ end, were dissolved in the printing buffer (150 mM sodium phosphate pH 8.5, 0.01% Sucrose monolaurate) to a concentration of 10 μM and printed by a piezoelectric spotter, SciFLEX ARRAYER S12 (Scienion, Berlin, Germany) on silicon/silicon oxide slides coated with MCP-4 according to the protocol provided by the manufacturer. The spotting was performed at 20° C in an atmosphere of 60% humidity. After the spotting step the chips were incubated overnight and all residual reactive groups of the coating polymer were blocked as previously described (23).

### Microarray hybridization and detection

The products of the PCR reactions were mixed with 2.5 μL of 20X SSC hybridization buffer and brought to the final volume (25 μL) with H_2_O. The mixtures were heated at 95^°^C for 5 min to denature the DNA double strand. The solution was quickly centrifuged and chilled on ice for 1 min then it was spread onto the microarray. A cover slip (large enough to cover the entire spotted surface) was carefully placed on the microarray to avoid any bubble captured in. The slides were incubated in a sealed humid hybridization chamber at room temperature for 15 min. The hybridized silicon chips were then removed from the hybridization chamber and soaked briefly in 4X SSC buffer to remove the cover slips. Finally, the chips were washed at room temperature with 0.2X SSC for 1 min and 0.1X SSC for 1 min and then dried with a nitrogen flow. The hybridized silicon chips were scanned with InnoScan 710 (Innopsys, Carbonne, France). A green laser (λ_ex_ 532 nm) for the Cy3 dye was applied. The photomultiplier (PMT) tube gain and the laser power changed between different experiments. 16-bit TIFF images were analysed at 5 µm resolution. Data intensities were extracted with the Mapix software and the data analysis was performed for each experiment.

## Results and discussion

### Detection of SARS-CoV-2 nucleic acid from clinical nasopharyngeal swab samples with CovidArray

An oligonucleotide microarray targeting two regions (N1 and N2) of the Sars-CoV-2 nucleocapsid, and the human *RPP30*, was developed (Figure 1A). To interpret the results, the indications of the US CDC qPCR test were followed. In this assay, the fluorescence of capture spots indicates positivity. Briefly, a specimen is considered positive for SARS-CoV-2 if the two SARS-CoV-2 markers (N1, N2) produce a fluorescence signal that exceeds more than 3 times the standard deviation the signal of the no-template control (NTC). On the contrary, a specimen is considered negative if the SARS-CoV-2 markers (N1, N2) show a signal non discernible from that of the NTC. The *RPP30* gene, in a positive sample, may or may not be positive. It is possible that some samples may fail to exhibit RPP30 fluorescence due to low cell numbers in the original clinical sample. A negative RPP30 signal does not preclude the presence of Sars-CoV-2 virus RNA in a clinical specimen. On the other hand, the absence of the *RPP30* signal in a negative specimen makes the result invalid because the presence of a *RPP30* signal in a sample negative for N1 and N2 confirms the correct extraction of RNA.

**Figure 1.**
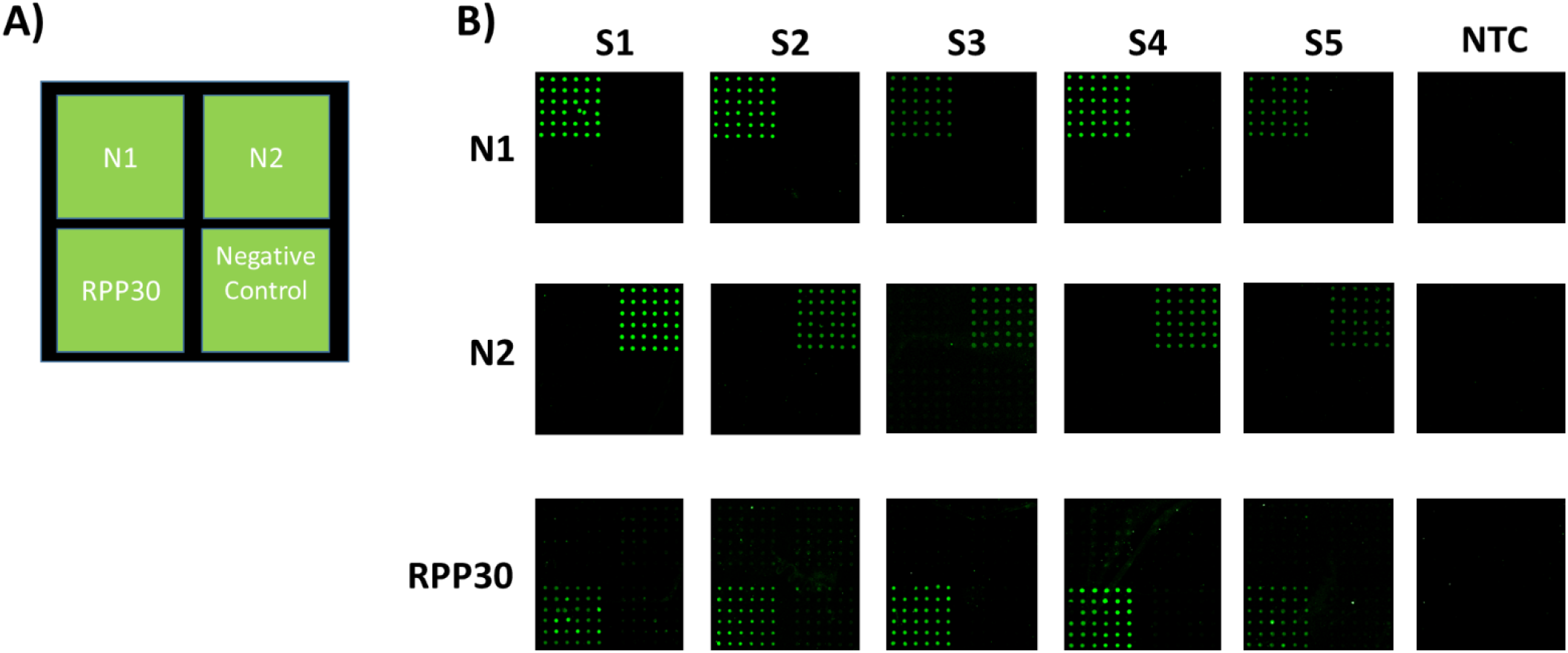
A) Spotting schema of the CovidArray. N1 and N2: nucleocapsid1 and 2 regions; *RPP30*: human Ribonuclease P gene; negative control: not correlated oligonucleotide probe. B) Cy3 fluorescence images of eighteen different silicon chips hybridized (from top to bottom) with N1, N2, and RPP30 amplicons of five nasopharyngeal samples (S1-S5) and with the NTC. Laser Power: Low; PMT: 5%

In this work, to speed up the test, the viral RNA was extracted from the nasopharyngeal swabs (stored at -80°C) using RNAGEM kit. To optimize the assay and reduce the analysis time, the transcripts were amplified at different number of cycles (data not show). The optimization showed that the array is able to detect, after only 25 cycles, amplicons that are detectable at higher number of cycles with standard RT-qPCR. A decrease of the number of cycles to 25 led to reduction of the analysis time (1h of reaction).

Firstly, it was demonstrated the correct assignment of samples previously assayed in the laboratory of Immunology at University of Milan (L. Sacco Hospital) with standard qPCR technology. In particular, five nasopharyngeal swabs (S1-S5) were analysed, four of which positive for the target N1 and N2 by RT-qPCR and one negative for both (S3). The four positive samples presented different viral loads resulting in different threshold cycles (Ct) in qPCR. In particular, for N1, the sample S1 crossed the threshold line at 20.00 cycles due to its high viral load; also sample S2 has a high viral load (22.00 cycles) while S4 and S5 have lower viral load and are detected at 38 and 36 cycles respectively (Table 1).

**Table1.** RT-qPCR for N1 and N2.

| Nasopharyngeal swabs | N1 qPCR (Ct)<sup>a</sup> | N2 qPCR (Ct)<sup>a</sup> |
| --- | --- | --- |
| S1 | 20.61 | 20.45 |
| S2 | 21.83 | 29.31 |
| S3 | N/A<sup>b</sup> | N/A<sup>b</sup> |
| S4 | 38.49 | N/A<sup>b</sup> |
| S5 | 35.86 | N/A<sup>b</sup> |

<sup>a</sup> Cycle Threshold; <sup>b</sup> not applicable

The Figure 1B shows the results of the microarray analysis of the swab samples and the control NTCs for the SARS-CoV-2 markers (N1 and N2) and for the RPP30 positive control. Amplicons of N1, N2 and *RPP30* were separately incubated on microarray chips. Three different silicon substrates were used to analyze one swab sample. As shown in Figure 1B, the fluorescence signal appears only at the location where the immobilized capture probe is complementary to the labelled PCR with no cross-hybridization and a good reproducibility from spot to spot. The absence of fluorescence in the NTC subarrays is essential as it allows discriminating low-signal samples from background signals. The same samples were also assayed by ddPCR. Indeed, one of our samples (S3) from a patient negative according to the RT-qPCR assay, but with symptoms attributable to Covid-19, was found to be positive by CovidArray in agreement with ddPCR (N1=34.5 copies/µL; N2= 7.6 copies/µL). CovidArray matches the sensitivity of ddPCR (Figure 2).

**Figure 2.**
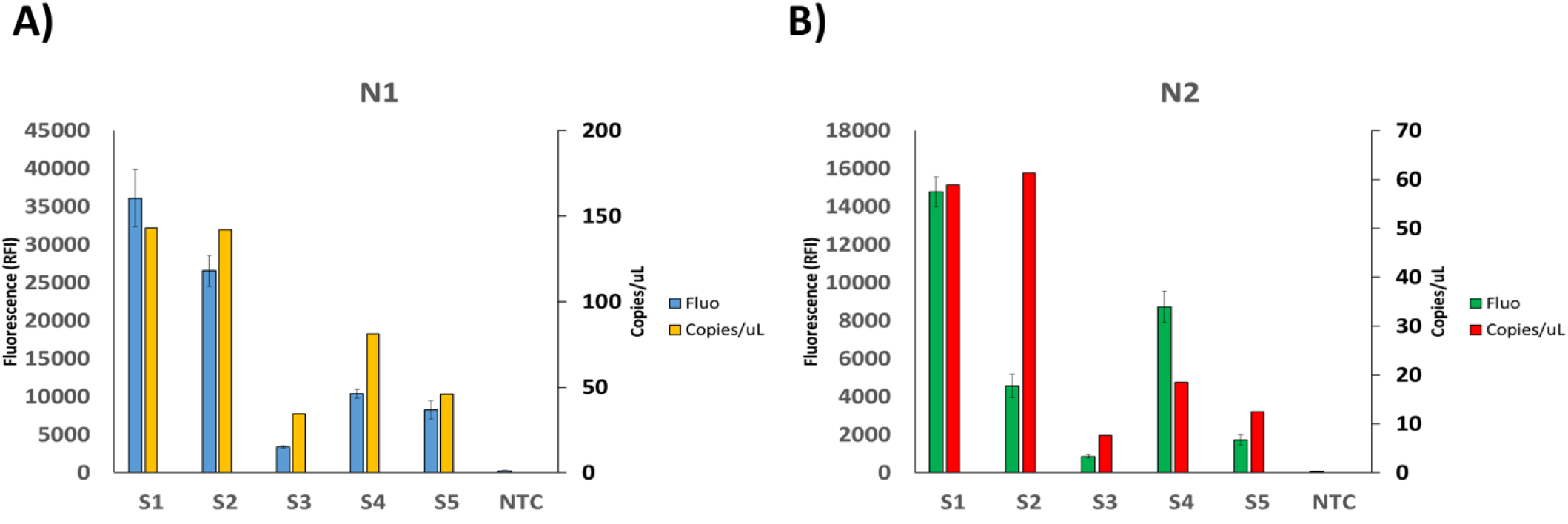
Plots showing the relative fluorescence intensity (blue and green bars) of the images in Figure 1B, and the copies number/μL of the corresponding ddPCR (yellow and red bars), for the N1 (A) and N2 (B) markers.

The sensitivity increase of the proposed method reduces the number of false-negative results. It also shortens the analysis time by reducing the number of PCR cycles required to detect a positivity. The fluorescence signal intensity of the spots in CovidArray correlates well with the viral load of the five samples with the higher fluorescence corresponding to the samples with the highest viral load. The CovidArray was further validated with seven nasopharyngeal swabs from IRCCS Mondino Foundation (Pavia), previously subjected to solid-phase extraction and RT-qPCR as reported in the experimental/materials and methods section. A 100% agreement between the two methods was found (Table 2). The fluorescence images for samples B243, N053 and NTC are shown in Supplemental Figure 1.

**Table2.**
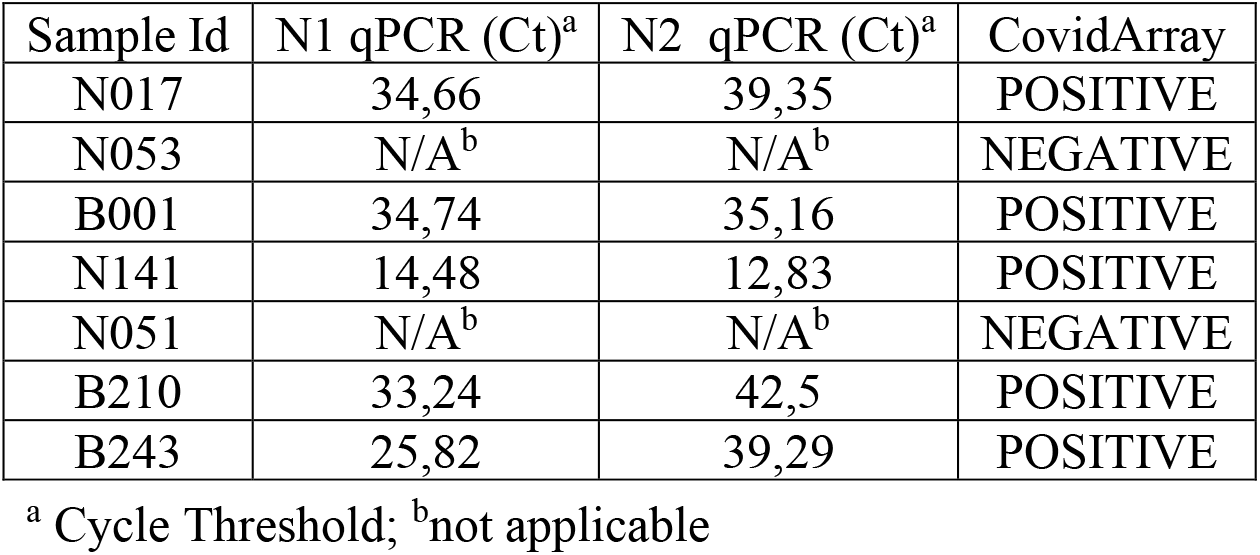
Correspondence between RT-qPCR Ct and CovidArray.

### Detection limit of CovidArray

Since the new SARS-CoV-2 emerged, researchers have struggled to develop highly sensitive molecular techniques to diagnose positive Covid-19 subjects effectively. According to the WHO and the Centre for Disease Control and Prevention (CDC), the gold standard for the diagnosis is qPCR. However, many studies have highlighted the presence of false negative results in RT-qPCR (24,25). Therefore, it is worthy to build-up novel robust methodologies that ensure high sensitivity useful not only for diagnostic purposes but also for the follow-up of patients and for monitoring of the viral load. To evaluate our method’s sensitivity, serial dilutions of linear DNA standard 2019-CoV Plasmid Control were tested using primer sets targeting N1 and N2 regions. The plasmid DNA was diluted to 50, 25, 5, 2.5, 0.5, 0.25, 0.05 copies/μL prior to undergo 25 cycles of PCR. To build the calibration curves for the two viral regions, the capture probes in 36 replicates (6×6 subarrays) were spotted on different coated silicon chips (one chip for each plasmid concentration). Samples with decreasing concentration of plasmid DNA were amplified, denatured, and finally hybridized for 15 minutes at room temperature. The value of fluorescence intensity detected for each of the seven plasmid concentrations together with the background fluorescence of the control sample with no plasmid DNA were plotted versus the number of copies of the plasmid per μL in the starting solution. Figure 3A shows the calibration curves for the region N1 and N2, respectively. The LODs (lowest concentration of detectable plasmid DNA) extrapolated for each marker are reported in Figure 3B. The determination of LOD is based on the equation: 3.3 *σ*/*s* where *s* is the slope of calibration curve and *σ* is the standard deviation of fluorescence background in the controlsample. The LODs found with this system are 1.16 copies/μL for the N1 and 0.81 copies/μL for the N2, respectively. These LODs are comparable with those declared by the various manufacturers of kits for qPCR with the difference that the number of standard amplification cycles for those methods is 40 while, in our approach only 25 cycles are sufficient to detect the target genes.

**Figure 3.**
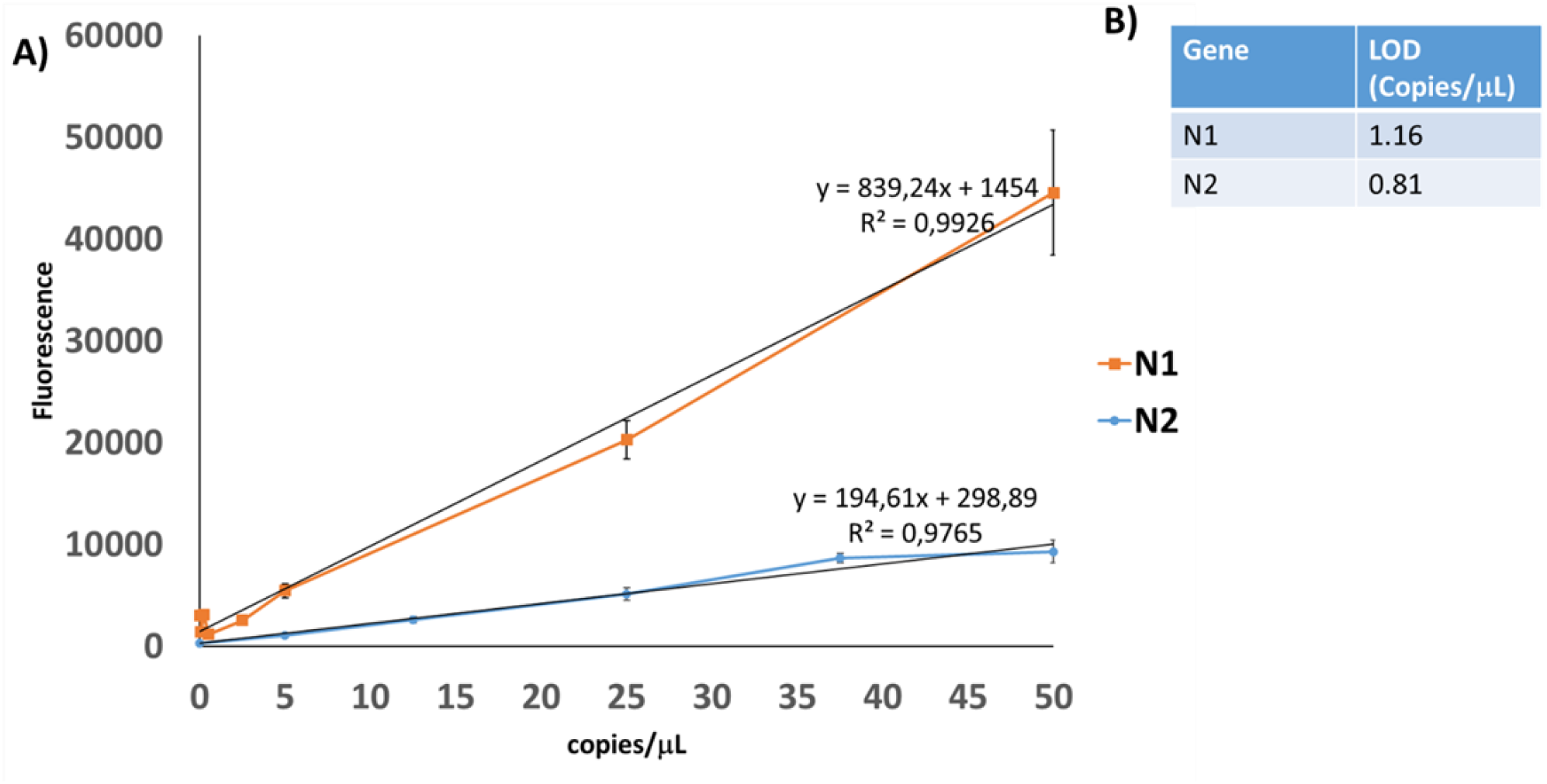
A) Plot of the relative fluorescence intensity of the N1 and N2 signals. The points correspond to the number of copies/μL of linear DNA standard 2019-CoV Plasmid Control. The value at point 0 represents the relative fluorescence intensity of the background with no 2019-CoV Plasmid Control. The equations of the trend lines are utilized to extrapolate the limit of detection (LOD) for the assay. B) The extrapolated limits of detection for the N1 and N2 marker.

### Multiplex capability of the CovidArray

One of the peculiar features of microarray technology is its multiplexing capability. Different genes can be revealed in a single hybridization assay by spotting onto the microarray substrate different capture probes specific to the target. In this work, we exploited the multiplexing capability of the CovidArray to detect the presence of the N1, N2 markers of SARS-CoV-2 and the RPP30 control gene on a single silicon chip in a single hybridization assay. We performed a triplex PCR, amplifying simultaneously the genes N1, N2 and *RPP30* using the cDNA produced by reverse transcription of the RNA extracted from the same 5 nasopharyngeal swabs as reported in the “Materials and Method” section. The triplex PCR was hybridized with the probes spotted on the same substrate. The simultaneous appearance of fluorescence signals on the subarrays corresponding to the N1 and N2 regions confirmed the positivity of the sample. The fluorescence signal of the RPP30 subarray was also detectable. In Figure 4B five triplex PCR’ hybridization results corresponding to the S1-S5 samples are shown. The NTC does not show significant fluorescence. In Figure 4C the histogram of fluorescence intensity indicates that the samples S1 and S2 have a higher fluorescence intensity confirming the higher viral load detected by the single-PCR CovidArray. Sample S3 which was considered negative by the RT-qPCR technique shows a weaker but detectable signal in agreement with ddPCR and single-PCR CovidArray.

**Figure 4.**
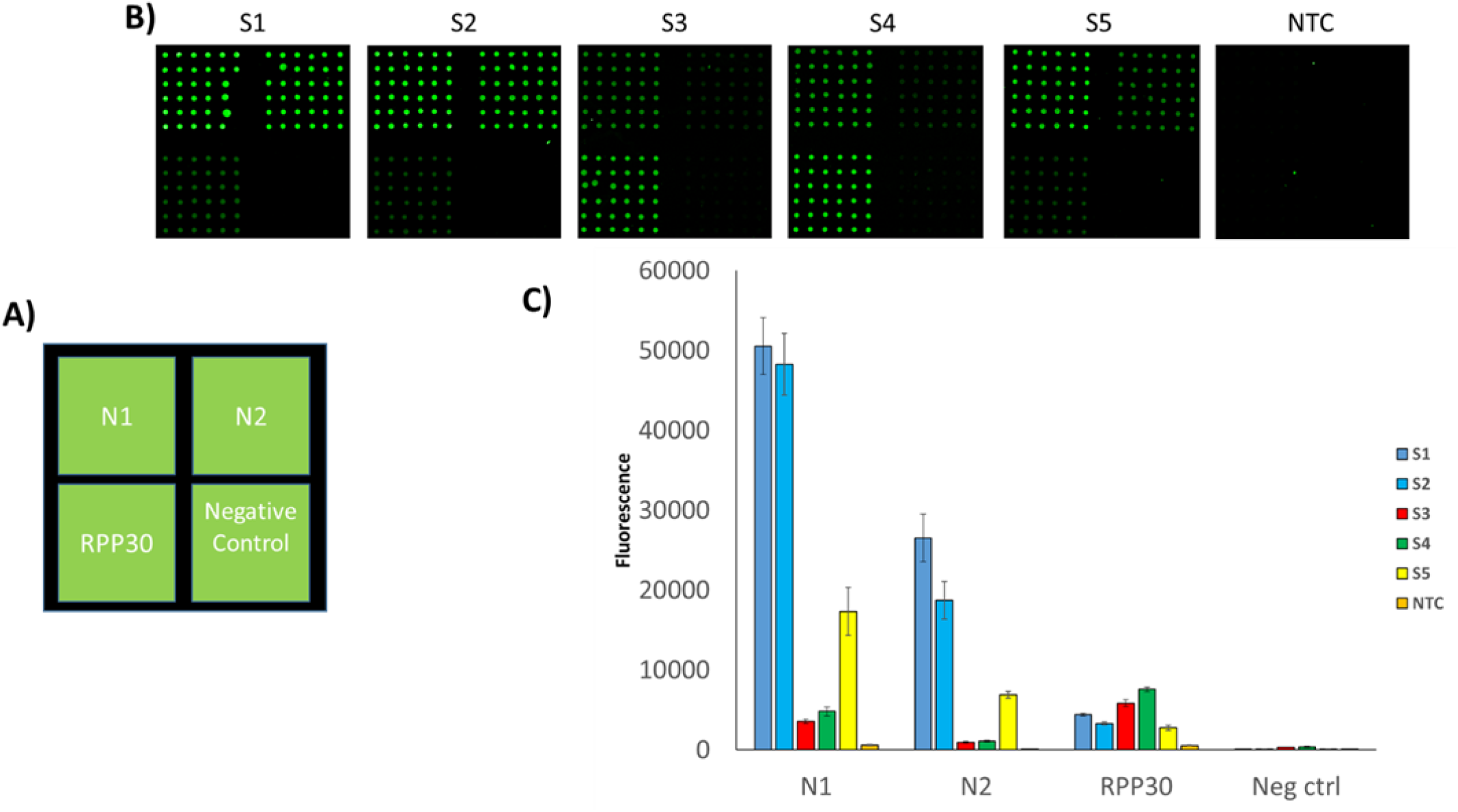
A) Spotting schema of the CovidArray. B) Cy3 fluorescence images of six silicon chips. Each CovidArray is hybridized with a triplex Cy3-labelled PCR specific for N1, N2, *RPP30* gene from the samples S1-S5 or with a No Template Control (NTC). C) The plot of the relative fluorescence intensity of the silicon chips shown in panel B. Laser Power: Low; PMT: 5%

## Conclusion

In summary, we describe a novel microarray platform, CovidArray, for the specific and sensitive detection of SARS-CoV-2 in nasopharyngeal swabs. This approach combines the physical-optical properties of the silicon substrate with the surface chemistry used to bind to the substrate the oligonucleotide capture probes specific to the virus’ genes to obtain a diagnostic tool of great sensitivity. In agreement with ddPCR, we correctly assigned 12 nasopharyngeal swabs of different origins. Thanks to the lower limit of detection of CovidArray, we identified a false-negative sample. Another feature of our system, also due to the high sensitivity of the CovidArray, is the decrease of the number of PCR cycles required to detect the viral markers which, in turn, leads to a significant reduction of the analysis time. A further contribution to speeding up the diagnostic assay is the use of an alternative method for extracting the viral RNA from clinical samples. In this work, we used an innovative approach, RNAGEM, commercially available by MicroGEM, to extract and purify viral RNA in less than 15 minutes. The total time required for the molecular test can thus range from about 3-6 hours of a standard process to about 2 hours with the CovidArray method. Moreover, RNAGEM provides an alternative to commercial RNA extraction kits that may undergo a shortage due to their massive use during the pandemic. Furthermore, in this work we have exploited the multiplexing capability of the microarray technology, to detect the presence of viral markers and the control sequence in a single assay. Finally, it is worth noticing the versatility of this approach. In fact, CovidArray could potentially allow differentiating SARS-CoV-2 from other viral and bacterial respiratory tract infections by merely adding new primers and capture probes to the same array, becoming a promising diagnostic tool suitable for routine diagnosis of a wide range of respiratory diseases.

## Data Availability

The data presented in this study are available on request from the
corresponding author

## List of abbreviations

Covid-19: CoronaVirus Disease 19
SARS-CoV-2: severe acute respiratory syndrome coronavirus 2
PCR: polymerase chain reaction
RT-qPCR: reverse transcription-quantitative polymerase chain reaction
qPCR: quantitative polymerase chain reaction
Copoly(DMA-NAS-MAPS): N,N-dimethylacrylamide (DMA), N,N-acryloyloxysuccinimide (NAS), and 3-(trimethoxysilyl) propyl methacrylate (MAPS) copolymer
ddPCR: droplet digital PCR
LOD: limit of detection
RPP30: Ribonuclease P gene
SSC: saline sodium citrate
SDS: sodium dodecyl sulfate
UTM: universal transport medium
PMT: photomultiplier tube

## Acknowledgments

We are grateful to Riccardo Vago (Università Vita-Salute S. Raffaele, Milan) and Greta Bergamaschi (SCITEC CNR, Milan) for their contribution without which this work would have been much more difficult. We also thank Giuseppina Sannino (MicroGEM UK Ltd, Southampton, United Kingdom) for her comments and for the supply of the RNAGEM Extraction Kit.

## Notes

### Competing Interest Statement

The authors have declared no competing interest.

### Funding Statement

This project has received funding from the Horizon 2020 research and
innovation programme of the European Union under grant agreement No 766466.

### Author Declarations

The protocol was approved by the local Medical Ethical and Institutional Review Board (Milan, area 1, #154082020). We obtained the informed consent from the patients, according to CARE guidelines and in compliance with the Declaration of Helsinki principles.

