## Supplemental data for "CovidArray: a microarray-based assay with high sensitivity for the detection of SARS-CoV-2 in nasopharyngeal swabs"

### ONLINE SUPPLEMENTAL DATA

#### Materials and Methods

#### Samples collection

Five nasopharyngeal swabs have been collected at the Unit of Obstetrics and Gynecology, L. Sacco COVID19-hub Hospital, ASST Fatebenefratelli Sacco, and seven nasopharyngeal swabs have been collected at the IRCCS Mondino Foundation (Pavia). This study was developed on existing samples collected during standard diagnostic tests that were positive to SARS-CoV-2 RNA detection. Subjects participating in the study gave their informed consent (oral or written) for SARS-CoV-2 analysis. The protocol was approved by the local Medical Ethical and Institutional Review Board (Milan, area 1, #154082020). We obtained the informed consent from the patients, according to CARE guidelines and in compliance with the Declaration of Helsinki principles.

#### RNA extraction and RT-qPCR

Maxwell® RSC Viral Total Nucleic Acid Purification Kit was used to extract RNA from 250 µL of the five nasopharyngeal swabs preservation media (from L. Sacco COVID19-hub Hospital) employing the Maxwell® RSC Instrument (Promega, Fitchburg, WI, USA) while RNAs from 350 µL of the seven nasopharyngeal swabs UTM (IRCCS Mondino Foundation) have been isolated by Magnetic bead method using an automatic nucleic acid purification system (GenePure Pro BIOER) for qPCR testing.

Dual labelled TaqMan probes with 5′-6-FAM fluorescent dye and 3′- BHQ-1 quencher for SARS-CoV-2 target sequences N1 was used for the detection of viral RNA. For internal reference control, a pair of primers and TaqMan probe for human Ribonuclease P gene (*RPP30*), labelled with 5′-HEX fluorescent dye and 3′-BHQ-1 quenchers were used. Primers use have been indicated by US Centers for Disease Control and Prevention [CDC 2019-Novel Coronavirus (2019-nCoV) Real-Time RT-PCR Diagnostic Panel].

For reaction mix, 3 µL of extracted RNA and 7 µL of Go-Script One-Step PCR mix (Promega, Madison, WI, USA) have been used for qPCR in CFX96 (BioRad, Richmond, CA) at Sacco Hospital, while 5 µL of extracted RNA, 5 µL of Reliance One-Step RT-qPCR Supermix (BioRad, Richmond, CA), 1 µL of RT enzyme and 4 µL of water have been used at Mondino Foundation, depending on the protocol used. Cycling conditions were 50°C for 10 min, 95°C for 3 min, followed by 40 cycles of amplification (95°C for 10 s and 60°C for 30 sec). qPCR analysis has been considered valid in all samples in which *RPP30* gene has been detected. Positive samples were determinate by Cycle threshold (Ct) of N1 and N2 gene minor of 40.

**Reverse Transcription**

8 µL of the RNA extracted were converted in cDNA using the SuperScript™ First-Strand Synthesis System (III) kit from ThermoFisher. The reverse transcription step was carried out according to the manufacturer's instructions. The obtained cDNA was diluted 1:20.

**droplet digital PCR (ddPCR)**

We employed the QX100™ Droplet Digital™ PCR System (Bio-Rad Laboratories, Hercules, CA, USA). 9 µL of cDNA previously diluted 1:20 were mixed with primers and fluorophore labeled commercial probes (2019-nCoV CDC EUA Authorized qPCR Probe Assay primer/probe mix, Integrated DNA Technologies) specific for the amplification of N1, N2 and *RPP30* genes as previously reported. The volume of the PCR mix was 20 µL including 10 µL of ddPCR™ Supermix for Probes (No dUTP) and 1 µL of primers/probe. The droplet emulsion was thermally cycled on C1000 Touch Thermal Cycler (Bio-Rad) instrument. Cycling conditions were 95°C for 5 min, followed by 40 cycles of amplification (94°C for 30 s and 60°C for 1 min), ending with 98°C for 10 min, according to the manufacturer's protocol. The copies of the target gene were calculated automatically by the QuantaSoft™ software version 1.7.4 (Bio-Rad).

**2019- CoV Plasmid Controls**

Plasmid controls contain the complete nucleocapsid gene from 2019-nCoV virus were provided by Integrated DNA Technologies and delivered at a concentration of 200,000 copies/ µL in IDTE pH 8.0.

**Supplemental data Table 1. Assay primer/probe sequences**

| Assay | Description | Oligonucleotide sequence | Amplicon size (bp) |
|---|---|---|---|
| N1 | Nucleocapside gene | For 5'-GACCCCAAAATCAGCGAAAT-3'<br>Rev[a] 5'-TCTGGTTACTGCCAGTTGAATCTG-3'<br>Probe[b] 5'-ACCCCGCATTACGTTTGGTGGACC-3' | 73 |
| N2 | Nucleocapside gene | For 5'-TTACAAACATTGGCCGCAAA-3'<br>Rev[a] 5'-GCGCGACATTCCGAAGAA-3'<br>Probe[b] 5'- ACAATTTGCCCCCAGCGCTTCAG-3' | 67 |
| *RPP30* | Ribonuclease P gene | For 5'-AGATTTGGACCTGCGAGCG-3'<br>Rev[a] 5'-GAGCGGCTGTCTCCACAAGT-3'<br>Probe[b] 5'-TTCTGACCTGAAGGCTCTGCGCG-3' | 65 |
| Negative control | Hybridization control | Probe[b] 5'-AGGGCTCTATTCAGCGTATT-3' | |

All the sequences are from the US CDC 2019-Novel Coronavirus (2019-nCoV) Real-Time RT-QPCR Diagnostic Panel. [a]The Reverse primers are labeled with Cyanine 3 in 5'-end. [b]The spotted capture probes are amino modified in 5'-end.

**Supplemental Figure**

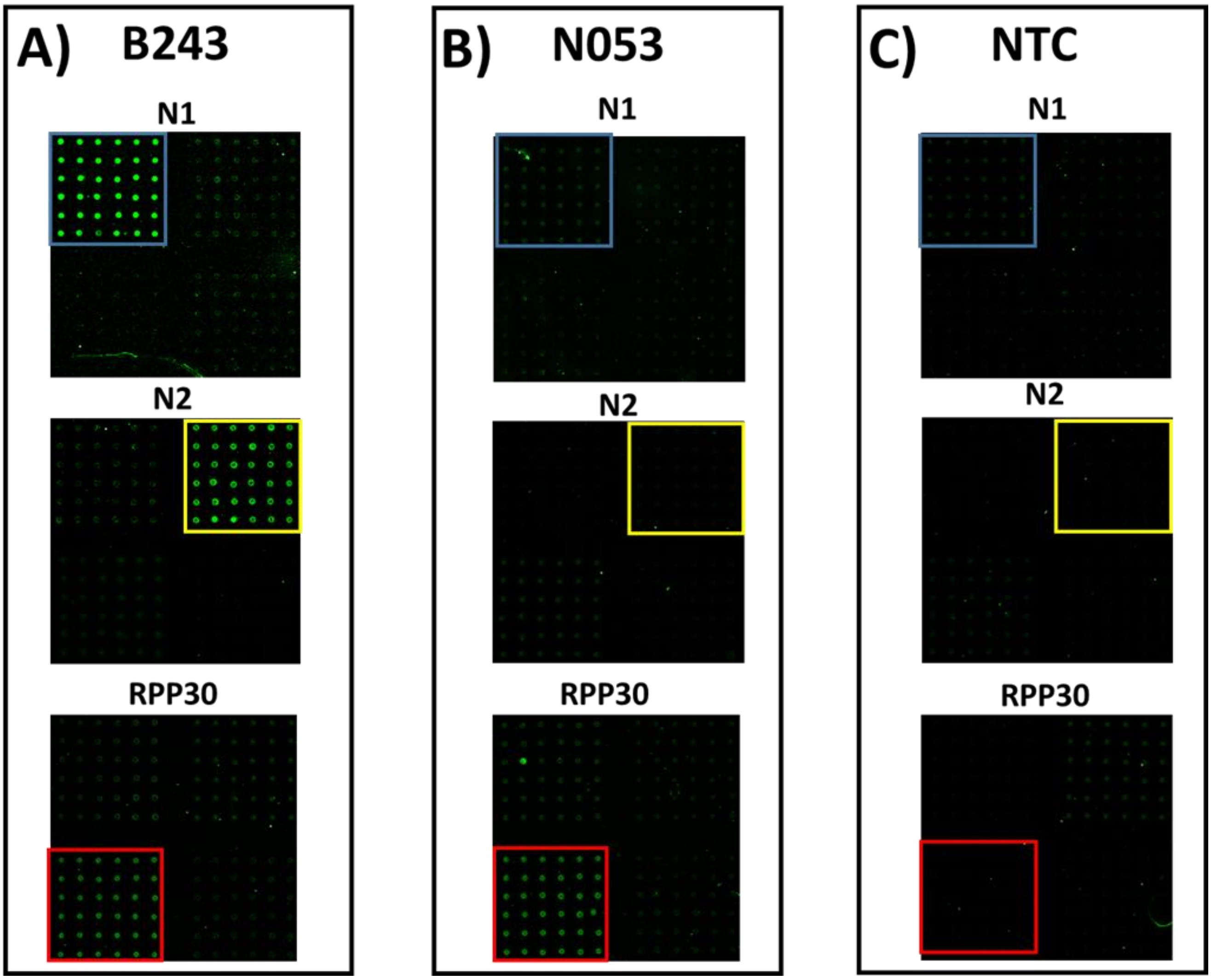


**Figure 1**

A) Cy3 fluorescence image of the CovidArray analysis of sample B243. N1, N2 and RPP30 subarrays are highlighted in blue, yellow, and red, respectively. B) Cy3 fluorescence images of the CovidArray analysis of sample N053. N1, N2 and RPP30 subarrays are highlighted in blue, yellow, and red, respectively. C) Cy3 fluorescence image of the CovidArray analysis of the No Template Control (NTC). N1, N2 and RPP30 subarrays are highlighted in blue, yellow, and red, respectively. Laser Power: Low; PMT: 5%
